# Modelling the potential role of super spreaders on COVID-19 transmission dynamics

**DOI:** 10.1101/2021.08.30.21262341

**Authors:** Josiah Mushanyu, Williams Chukwu, Farai Nyabadza, Gift Muchatibaya

## Abstract

Superspreading phenomenon has been observed in many infectious diseases and contributes significantly to public health burden in many countries. Superspreading events have recently been reported in the transmission of the COVID-19 pandemic. The present study uses a set of nine ordinary differential equations to investigate the impact of superspreading on COVID-19 dynamics. The model developed in this study addresses the heterogeineity in infectiousness by taking into account two forms of transmission rate functions for superspreaders based on clinical (infectivity level) and social or environmental (contact level). The basic reproduction number has been derived and the contribution of each infectious compartment towards the generation of new COVID-19 cases is ascertained. Data fitting was performed and parameter values were estimated within plausible ranges. Numerical simulations performed suggest that control measures that decrease the effective contact radius and increase the transmission rate exponent will be greatly beneficial in the control of COVID-19 in the presence of superspreading phenomena.

## 1 Introduction

Severe acute respiratory syndrome coronavirus 2 (SARS-COV-2) disease that originated in the Wuhan City of China is continuing to spread in most parts of the globe. COVID-19 has devastated many communities and countries and to date, it has resulted in more deaths than those caused by SARS and MERS combined. One of the major factor driving this pandemic is the super-spreading (SS) phenomenon. Wong et al. (2015) defined SS as an event that occurs when one individual spreads the pathogen to a disproportionate number of contacted individuals. The U.S. Center for Disease Control and Prevention categorised super spreaders as individuals that are responsible for greater than 10 secondary infections Mkhatshwa and Mummert (2010). For the case of SARS, a SS event was classified as one that occurs when an individual infects ≥ 8 individuals Shen et al. (2004). In a study investigating the impact of socio-environmental factors on TB transmission, Issarow et al. (2018) defined super spreaders as individuals exhaling ≥ 10 infectious respirable particles hr^*−*1^. However, concern has been raised about the largely perceived meaning of this term about the current COVID-19 pandemic especially in social media which has been viewed to be problematic Cave (1984). A call has been made to provide a more concise scientific definition that takes into account the behavioural, environmental and biological factors linked to SS to avoid stigmatisation of those infected thereby deterring potential control efforts of the pandemic Cave (1984). The need to counteract super spreading events becomes paramount considering that SS is associated with an exponential increase in the early stages of the infection and subsequent sustained community transmissions Frieden (2020); Wong et al. (2015); Lau et al. (2017); Walling and Teunis (2004); Yin et al. (2020).

SS has also been reported for other infectious diseases such as HIV, Typhoid, Ebola virus disease, gonorrhoea, tuberculosis (TB), measles, mumps and rubella, smallpox, monkey-pox, pneumonic plague, flu pandemic, SARS and MERS-COV disease Hui (2016); Stein (2011); Melsew et al. (2020); Frieden (2020); Mkhatshwa and Mummert (2010); Lloyd-smith et al. (2005). SS for typhoid was noted in the early 20th century where a cook Mary Mallon infected more than 50 individuals Kim et al. (2016.); Hui (2016); Stein (2011); Marineli et al. (2013). For the tuberculosis disease, SS was reported in a study by Riley et al. Riley et al. (1962) which showed that amongst many patients with smear-positive and cavitary tuberculosis only 3 out of 77 patients were responsible for 73% of tuberculosis infections. SS during the West Africa Ebola virus disease outbreak was reported to have sustained the epidemic with 3% of infected individuals associated with 61% of infections Lau et al. (2017). SS events were also reported in the spread of gonorrhoea where few individuals had higher infectivity as compared to others Hethcote and Yorke (1984). SS events have also been reported in the spread of animal infections in buffaloes, deer mice and cattle Maeno (2011); Mkhatshwa and Mummert (2010). SS has not been limited to the spread of infections only. In a study by Yin et al., Yin et al. (2020), the authors described some information super spreaders termed ‘opinion leaders’ who were responsible for the enormous propagation of important information on public health interventions for COVID-19 in Wuhan City, Hubei Province of China. The authors developed a mathematical model to examine the dynamics of information propagation in the Chinese Sina-microblog through the application of an opinion-leader susceptible-forwarding-immune (OL-SFI) model.

Information about the epidemiology of the COVID-19 disease is currently limited but there are reports of super spreading events linked to the disease Frieden (2020); Wang et al. (2020). In South Korea, one of the members (patient 31) of the Shincheonji church infected several other members resulting in an increased number of recorded deaths attributable to this church’s gathering kasulis (2020). A 70-year old preacher who later succumbed to the COVID-19 disease was suspected to have led to the death of around 30 people and quarantine of approximately 40, 000 individuals in the state of Punjab, India BBC news (2020). In Zimbabwe, some reports indicated that some escapees who were unaware of their COVID-19 status infected their household members. This prompted community members to report any suspected cases of escapees in their territory. A variety of factors thought to drive the SS phenomenon include environmental, host, pathogen, and behavioural factors Stein (2011). Understanding the specific impact of these factors can facilitate informed prevention and control strategies. The risk of SS differs across regions based on the community’s or country’s cultural and socio-economic attributes Frieden (2020).

Various mathematical techniques and tools have been used for studying the SS phenomenon in infectious diseases. These include, Ordinary differential equations Kemper (1980); Mkhatshwa and Mummert (2010); Ndairou et al. (2020), stochastic differential equations Maeno (2011), agent-based modelling (ABM) Yunhwan et al. (2018), negative binomial distribution Lloyd-smith et al. (2005), branching process models in discrete and continuous time generations James et al. (2007); Garske and Rhodes (2008), contact network models Aihaira et al. (2004); zhang et al. (2019). We describe the pioneer ordinary differential equations models developed to study SS events. Kemper (1980) formulated an *SI*_1_*I*_2_*R* model with *S*(*t*) representing the susceptible population, *I*_1_(*t*) representing the first class of infectives, *I*_2_(*t*) representing the second class of infectives and *R*(*t*) representing the class of the removed individuals. It was suggested that this model can be suitable to model SS events by assuming different transmission rates *r*_1_ and *r*_2_ for *I*_1_ and *I*_2_ respectively, where the presence of SS events is captured by using higher than normal transmission rates. Mkhatshwa and Mummert (2010) modified the *SI*_1_*I*_2_*R* model in Kemper (1980) to come up with an *SIPR* model for the SARS outbreak where *S*(*t*) are the susceptibles, *I*(*t*) are the regularly infected individuals, *P*(*t*) are the super spreader individuals and *R*(*t*) are the recovered individuals. The *SIPR* model by Mkhatshwa and Mummert (2010) assumed the same transmission rate *β* for both the *I*(*t*) and *P* (*t*) classes but applied the 20*/*80 rule in Woolhouse et al. (1997) by apportioning *b* the probability that a newly infected will be regular and the remainder (1 − *b*) the probability that a newly infected will be a super spreader. Also, the authors captured SS events by assuming that the overall time spent outside quarantine and isolation for regularly infected individuals (*ν*_1_ days) is less than that of super spreader individuals (*ν*_2_ days), that is, *ν*_1_ *< ν*_2_. This prolonged time outside quarantine and isolation for a super spreader individual means that a superspreader will have increased chances to have many contacts with susceptibles as compared to the regularly infected individual. One of the limitations of the *SIPR* model involved the assumption of the same transmission rate for both the regularly infected individuals and the super spreader individuals.

In this paper, we extend the work done on modelling the SS phenomenon. We adopt the transmission rate functions developed for the agent-based model in Yunhwan et al. (2018) that provides a general mathematical tool for outbreaks with SS events. The model developed in this study takes into account the heterogeneity of transmission among infected individuals not considered in previous SS models. Heterogeneity in infectiousness has been classified in two forms namely; clinical (infectivity level) and social or environmental (contact level) Yunhwan et al. (2018). Understanding the role of these forms of transmission will be beneficial in informing public health policy. The model also takes into account the most effective documented control measures such as early discovery through contact tracing and diagnosis followed by intervention in the form of isolation and treatment Wong et al. (2015). Inclusion of such important control measures in our model will enable assessment of their relative contribution in curtailing SS.

The manuscript is organised as follows: In Section 2, a mathematical model that describes the effect of super-spreaders on the COVID-19 pandemic is formulated and its mathematical analysis given in Section 3. In Section 4, we present the numerical simulations confirming some of the analytical results. Finally, a brief discussion is given in Section 5.

## 2 Model formulation

To study the spread of COVID-19 driven by SS, we consider the human population sub-divided into nine distinct compartments representing different infection statuses with respect to the disease, (see Table 1 for the description of the state variables and parameters). We consider transmission rates for normal or regular spreaders (*λ*_*n*_(*t*)) and super spreaders 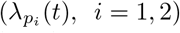. We take into account two forms of transmission rates for super spreaders based on infectivity level and frequency of contacts. The transmission rate for regular spreaders is given by

**Table 1.** Model state variables and parameters.

| State variable | Description |
| --- | --- |
| $S$ | Individuals at risk of being infected (Susceptibles) |
| $E$ | Individuals exposed to COVID-19 but not infectious (Exposed) |
| $A$ | Regularly infected asymptomatic individuals |
| $I$ | Regularly infected symptomatic individuals |
| $P_1$ | Super-spreaders with higher infectivity levels than regular spreaders |
| $P_2$ | Super-spreaders with more contacts than regular spreaders |
| $Q$ | Quarantined/hospitalised individuals |
| $R$ | Recovered individuals |
| $D$ | Deceased individuals |
| Symbol | Parameter description |
| $r_0$ | Effective contact radius |
| $b$ | Transmission rate exponent |
| $\theta$ | Longer effective contact radius |
| $\varepsilon$ | Role of governmental action |
| $\beta_0$ | Transmission rate parameter |
| $\eta_1$ | Modification parameter for $A$ individuals |
| $\eta_2$ | Modification parameter for $Q$ individuals |
| $\sigma^{-1}$ | Incubation period |
| $\rho^{-1}$ | Infectious period for individuals in class $I$ |
| $\gamma^{-1}$ | Infectious period for individuals in class $A$ |
| $\delta^{-1}$ | Infectious period for individuals in class $P_1$ |
| $\alpha^{-1}$ | Infectious period for individuals in class $P_2$ |
| $m_1$ | Proportion of individuals in class $I$ who recover |
| $m_2$ | Proportion of individuals in class $I$ who are quarantined |
| $f$ | Proportion of individuals in class $A$ who are quarantined |
| $q_1$ | Proportion of individuals in class $P_1$ who recover |
| $q_2$ | Proportion of individuals in class $P_1$ who are quarantined |
| $h_1$ | Proportion of individuals in class $P_2$ who recover |
| $h_2$ | Proportion of individuals in class $P_2$ who are quarantined |
| $\phi^{-1}$ | Average period one stays in quarantine |
| $g$ | Proportion of individuals in class $Q$ who recover |
| $r$ | Distance between individuals in the $I$, $A$ or $Q$ status and the $S$ individuals |
| $k_1$ | Proportion of $E$ individuals joining the $I$ class at the end of the incubation period |
| $k_2$ | Proportion of $E$ individuals joining the $A$ class at the end of the incubation period |
| $k_3$ | Proportion of $E$ individuals joining the $P_2$ class at the end of the incubation period |

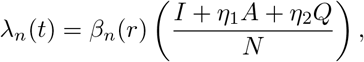

Where 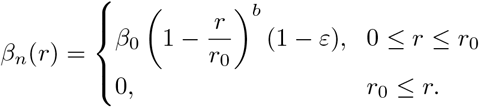

Here, *r*_0_ is the effective contact radius and *b* is the exponent satisfying *b >* 1. Take note that *β*_*n*_(*r*) is a decreasing function. We define *r* as the distance between individuals in the *I, A* or *Q* status and the susceptible individuals *S*. The parameter *ε* models the role of governmental action that includes use of sanitisers, wearing of face masks, social distancing, lockdowns etc.

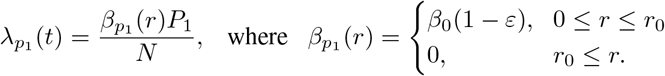

We now define the transmission rate function for super spreaders class *P*_1_ who are assumed to have higher infectivity level as follows:

Take note that for this case 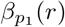 takes the form of a constant function with respect to the distance *r*.

We now define the transmission rate function for super spreaders class *P*_2_ who are assumed to have more contacts than regular spreaders as follows:

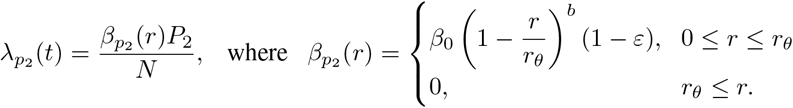

The transmission rate function for super spreaders class *P*_2_ is modelled by considering a longer effective contact radius where *r*_*θ*_ = *θr*_0_ with *θ >* 1. This longer effective contact radius can be regarded as an equivalent of staying outside quarantine and isolation for a longer time as compared to normal or regular spreaders.

The schematic diagram is given below. The system of non-linear ordinary differential equations is obtained upon combining details given in the model description above together with model flow diagram (Figure 1), as follows:

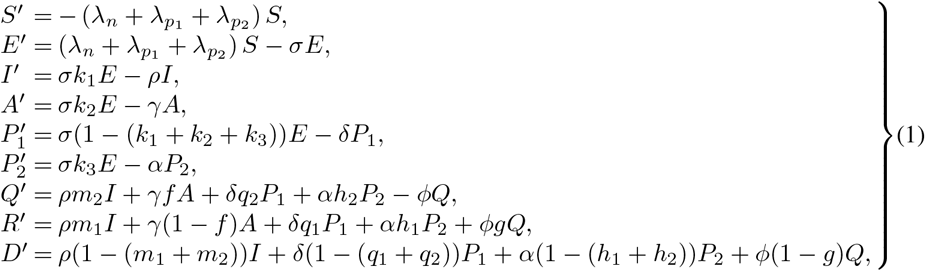

with the initial conditions

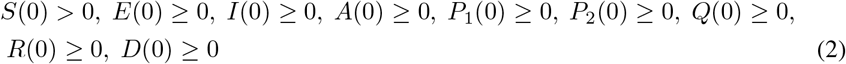

where all the model parameters are considered to be non-negative.

**Figure 1.**
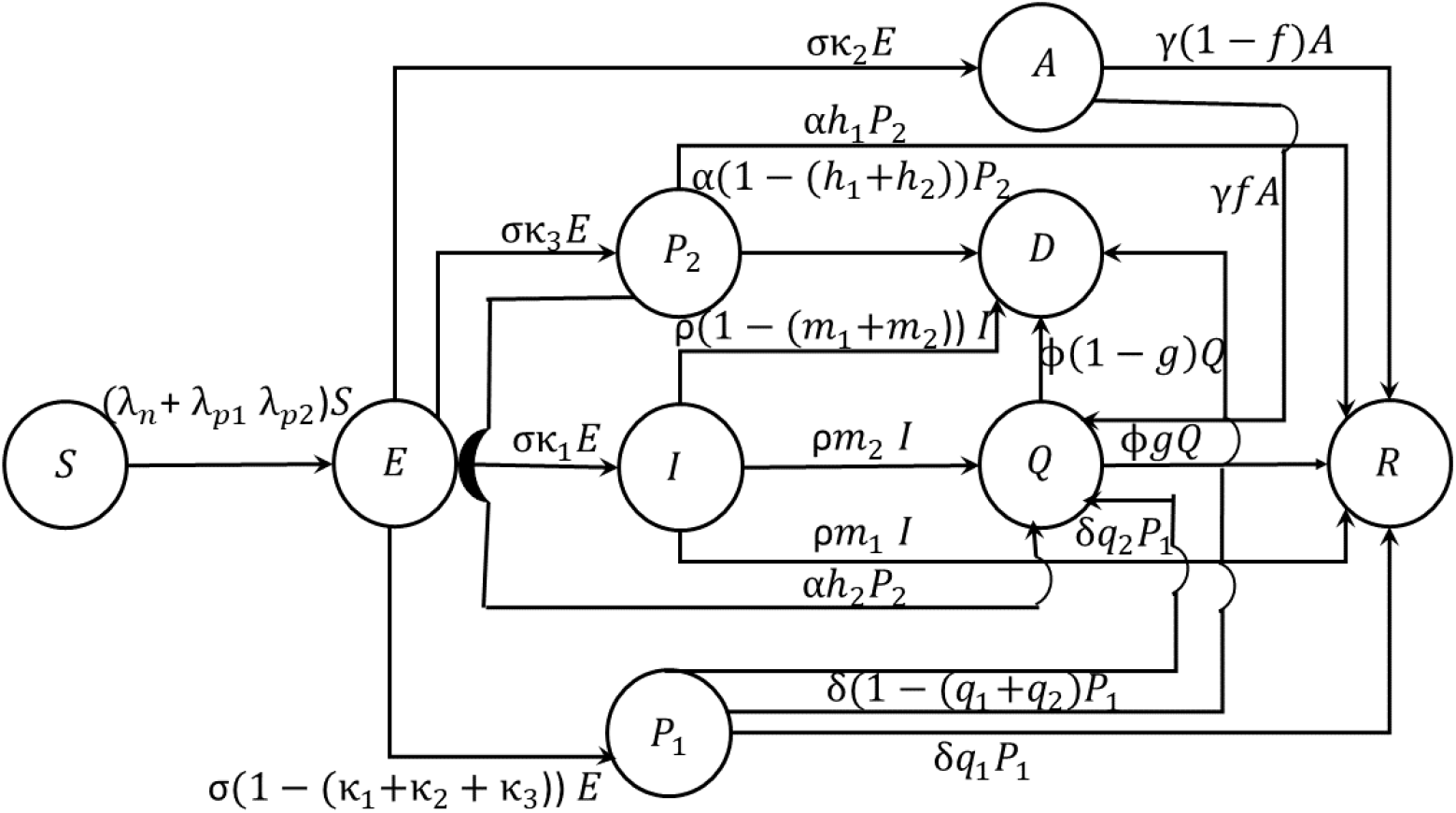
Model flow diagram.

**Figure 2.**
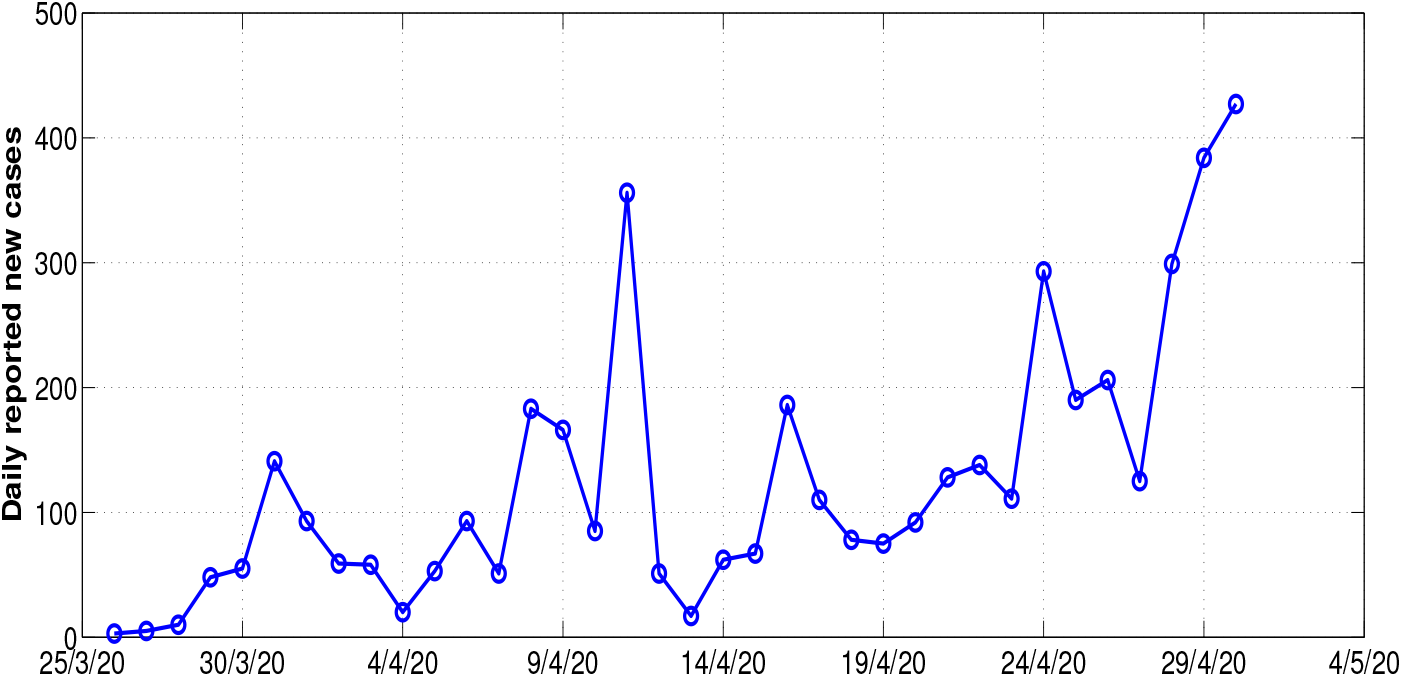
Daily reported new cases in Delhi starting on the 25th of March 2020 and ending on the 29th of April 2020.

**Figure 3.**
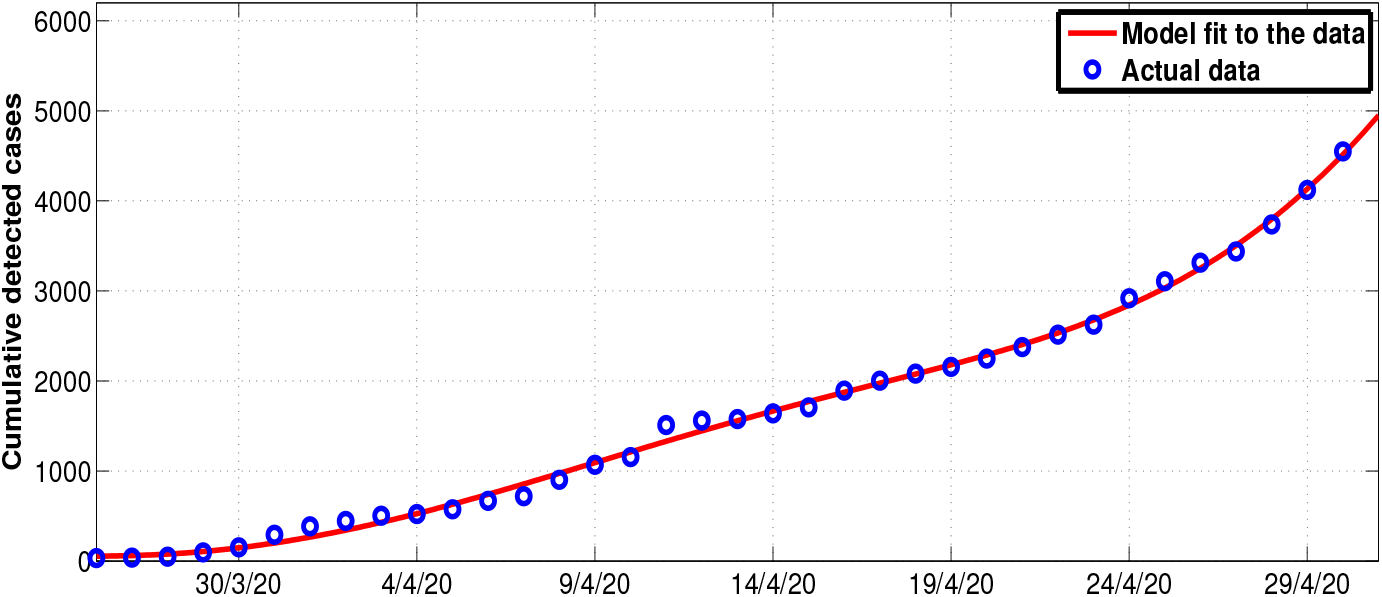
System (1) fitted to data for cumulative COVID-19 cases in Delhi starting from the 25th of March 2020 and ending on the 29th of April 2020. See Table 2. This period covers the whole of the 1st lockdown period which was later on extended. The *blue circles* indicate the actual data and the *solid red line* indicates the model fit to the data.

## 3 Mathematical analysis

### 3.1 Invariant region

Let Γ be the biologically meaningful invariant region for system (1) which is contained in 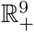. We prove that the solutions of the system exist and are bounded. This is ascertained by using the following theorem on the existence of solutions stated as follows.

#### Theorem 1

*The feasible region defined by* 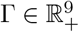, *given as*

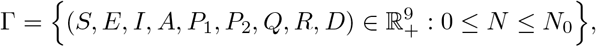

*is positively invariant with respect to system (1) with initial conditions (2)*.

*Proof*: The total change in human population obtained by summing all the nine equations of system (1) gives a constant population

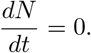

Thus, the lim_*t*→∞_ sup *N* ≤ *N*_0_. Clearly, we see that Γ is positively invariant with respect to system (1). Hence, all the solutions of system (1) exist, and are biologically meaningful, bounded, and remain in Γ for *t* ≥ 0. □

### 3.2 Positivity of solutions

We show that the solutions of system (1) are positive subject to the following initial conditions 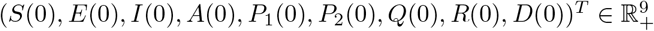. To ascertain this, we use Lemma 1 as in Yang et al. (1996).

#### Theorem 2

*Each solution* (*S*(*t*), *E*(*t*), *I*(*t*), *A*(*t*), *P*_1_(*t*), *P*_2_(*t*), *Q*(*t*), *R*(*t*), *D*(*t*)) *of system (1) with positive initial values (2) is positive for all t* ≥ 0.

*Proof*: Applying Lemma 1 as stated in Yang et al. (1996) we let

*X* = (*S, E, I, A, P*_1_, *P*_2_, *Q, R, D*)^*T*^ and *g*(*X*) = (*g*_1_(*X*), *g*_2_(*X*), …, *g*_9_(*X*))^*T*^ and re-write system (1) as follows

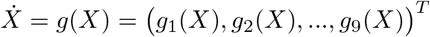

where

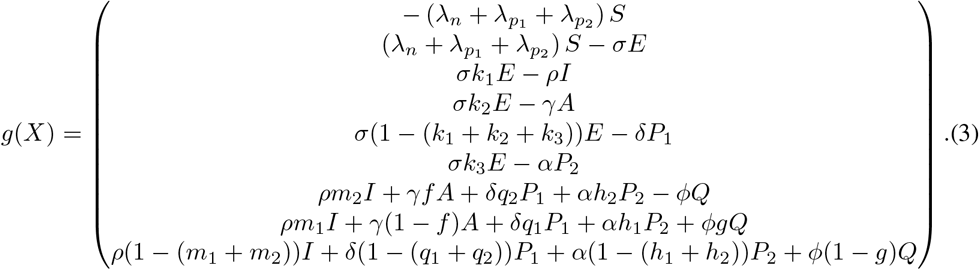

We determine if equation (3) satisfies the conditions given by Lemma 1 (see Yang et al. (1996)) by equating all the state variables for each class to zero which gives:

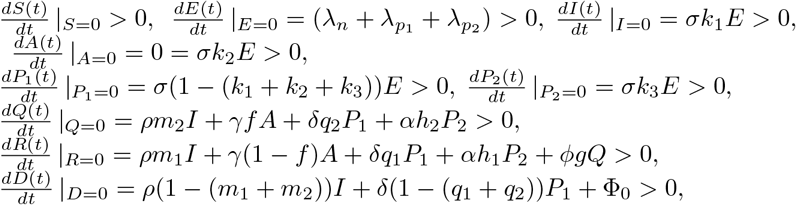

in which Φ_0_ = *α*(1 − (*h*_1_ + *h*_2_))*P*_2_ + *ϕ*(1 − *g*)*Q*. Thus, we have that the solutions of system (1) are positive. □

### 3.3 The model reproduction number and the steady states

Employing the next generation matrix approach by van den Driessche and Watmough van Driessche and Watmough (2002) we obtain

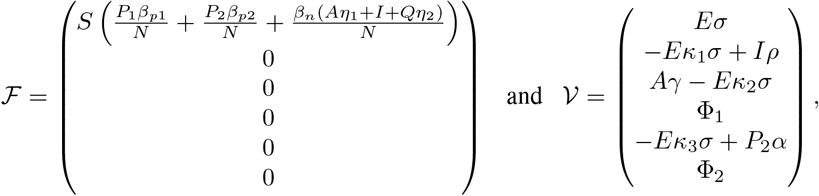

in which Φ_1_ = −*Eσ* (−*κ*_1_ −*κ*_2_ −*κ*_3_ + 1) + *P*_1_*δ* and

Φ_2_ = −*Afγ* −*Im*_2_*ρ* − *P*_1_*δq*_2_ − *P*_2_*αh*_2_ + *Qϕ*. Computing the Jacobian of *ℱ* and *𝒱* at the disease-free equilibrium leads to

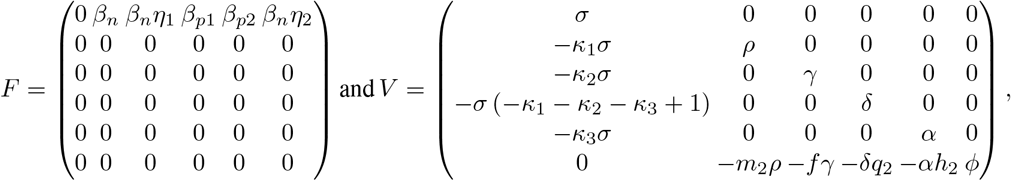

where the model basic reproduction number is the given as *ℛ*_0_ = *ρ*(*FV* ^*−*1^), with

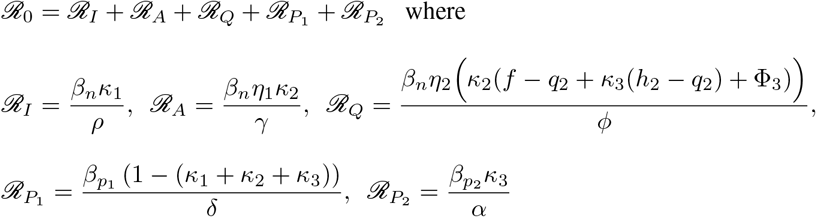

where Φ_3_ = *κ*_1_(*m*_2_ − *q*_2_) + *q*_2_. Here, 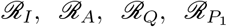 and 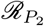 represent the contributions of classes *I, A, Q, P*_1_ and *P*_2_ respectively.

## 4 Numerical simulations

### 4.1 Parameter estimation

In order to estimate model parameters, we use data from a high population density country where SS has been reported as a result of high human to human social contact rate. We use the data for Delhi, one of the most affected provinces in India as a result of the Tablighi Jamaat event which took place from the 1st of March 2020 up to the 21st of March 2020. This event has since been regarded as a superspreader event since many coronavirus cases across India have been attributed to attendees of this event who tested positive for coronavirus Wikipedia (2020). As of 31st of March 2020, India had about 24 people who tested positive among the SS attendees, 300 showed symptoms and 16 were quarantined Wikipedia (2020). According to the Union Health Ministry of India, as of the 18th of April 2020 the total number of confirmed cases were 4291, translating to 29.8% of India’s total number of cases. These were all linked to the SS event which took place from the 1st of March 2020 up to the 21st of March 2020 Indiatimes (2020).

We tabulate Delhi’s COVID-19 cumulative cases (Table 2). The data used here was taken from Delhidata (2020). We used the data from within the time period 25 March 2020 up to 3 May 2020. This period covers the initial 21 day lockdown instituted on the 25th of March 2020 and later extended to the 17th of May 2020 Biswas et al. (2020). Thus, the choice of these dates is because the impact of the disease spread was felt in India immediately after the SS event which ended on the 21st of March 2020.

**Table 2.** Confirmed COVID-19 cases in Delhi.

|  |  |  |  |  |  |  |  |  |
| --- | --- | --- | --- | --- | --- | --- | --- | --- |
| **Date** | 25/3/20 | 26/3/20 | 27/3/20 | 28/3/20 | 29/3/20 | 30/3/20 | 31/3/20 | 1/4/20 |
| **Total cases** | 34 | 39 | 49 | 97 | 152 | 293 | 386 | 445 |
| **Date** | 2/4/20 | 3/4/20 | 4/4/20 | 5/4/20 | 6/4/20 | 7/4/20 | 8/4/20 | 9/4/20 |
| **Total cases** | 503 | 523 | 576 | 669 | 720 | 903 | 1069 | 1154 |
| **Date** | 10/4/20 | 11/4/20 | 12/4/20 | 13/4/20 | 14/4/20 | 15/4/20 | 16/4/20 | 17/4/20 |
| **Total cases** | 1510 | 1561 | 1578 | 1640 | 1707 | 1893 | 2003 | 2081 |
| **Date** | 18/4/20 | 19/4/20 | 20/4/20 | 21/4/20 | 22/4/20 | 23/4/20 | 24/4/20 | 25/4/20 |
| **Total cases** | 2156 | 2248 | 2376 | 2514 | 2625 | 2918 | 3108 | 3314 |
| **Date** | 26/4/20 | 27/4/20 | 28/4/20 | 29/4/20 |  |  |  |  |
| **Total cases** | 3439 | 3738 | 4122 | 4549 |  |  |  |  |

### 4.2 Model fitting

We fit system (1) to epidemiological data for Delhi from the 25th of March 2020 up to the 29th of April 2020. We make use of the Least Square (LS) fitting method which is a mathematical regression analysis tool used in determining the best fit for any given data set. The LS method provides the overall rationale for the placement of the best fit on the data set under study by giving the smallest sum of squares know as the variance Balcha (2020). One of LS’s advantages is that it allows modelers to estimate unknown parameter values by giving a lower bound and an upper bound from which the set of estimated parameter values that produce the best fit is obtained. Thus, in this work, we validate system (1) by fitting it to the data in Table 2 using the LS regression technique. This technique minimises the sum of the squared residuals given by:

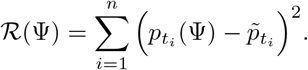

Here, Ψ represents all the model parameters in Table 3, *n* is the total number of data points used for the fitting process, 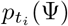 and 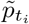 are the cumulative COVID-19 cases of infected individuals by our model predictions and the actual reported data for the time frame under investigation respectively.

**Table 3.** Parameter values obtained from data fitting and literature used for numerical simulations.

| Symbol | Base line value | Range | Source |
| --- | --- | --- | --- |
| $r_0$ | $\sqrt{6}$ | $0 - \sqrt{9}$ | Fujie et al. (2007) |
| $b$ | 2 | $1 - 10$ | Fujie et al. (2007) |
| $r$ | 0.2673 | $0 - 2$ | Data fit |
| $\theta$ | 8.3208 | $3 - 9$ | Data fit |
| $\varepsilon$ | 0.48576 | $0 - 1$ | Biswas et al. (2020) |
| $\beta_0$ | 0.88689 | $0 - 2$ | Biswas et al. (2020) |
| $\eta_1$ | 0.67047 | $0 - 1$ | Biswas et al. (2020) |
| $\eta_2$ | 0.06 | $0 - 1$ | Biswas et al. (2020) |
| $\sigma^{-1}$ | 5 days | $3 - 5$ | Lin et al. (2020) |
| $k_1$ | 0.580 | $0 - 1$ | Ndairou et al. (2020) |
| $k_2$ | 0.1321 | $0 - 1$ | Data fit |
| $k_3$ | 0.001 | $0 - 1$ | Ndairou et al. (2020) |
| $\rho^{-1}$ | 5 days | $1 - 18$ | Lin et al. (2020) |
| $\gamma^{-1}$ | 3 days | $1 - 10$ | Data fit |
| $\delta^{-1}$ | 10 days | $1 - 10$ | Data fit |
| $\alpha^{-1}$ | 7 days | $1 - 10$ | Data fit |
| $m_1$ | 0.7888 | $0 - 1$ | Data fit |
| $m_2$ | 0.19745 | $0 - 1$ | Data fit |
| $f$ | 0.00999 | $0 - 1$ | Data fit |
| $q_1$ | 0.0089 | $0 - 1$ | Data fit |
| $q_2$ | 0.0008796 | $0 - 1$ | Data fit |
| $h_1$ | 0.3254 | $0 - 1$ | Data fit |
| $h_2$ | 0.9859 | $0 - 1$ | Data fit |
| $\phi^{-1}$ | 7 days | $1 - 15$ | Data fit |
| $g$ | 0.7166 | $0 - 1$ | Data fit |

### 4.3 Numerical results

We use the estimated initial conditions for the exposed individuals *E*(0), regularly infected asymptomatic class *A*(0), the regularly infected symptomatic class *I*(0) and the quarantined/hospitalised individuals *Q*(0) in Biswas et al. (2020). The data used in Biswas *et al*. Biswas et al. (2020) was for the whole of India. We choose the estimated initial conditions from the second data set considered in Biswas et al. (2020) which covers the period 25th of March 2020 up to the 24th of April 2020 as this was observed to be the best estimator with least standard error unlike the first data set covering the period 1st of March 2020 up to the 24th of March 2020. In order to apply these initial conditions in the present study, we consider Delhi’s population as a fraction of the total population of India. Thus, we obtain the following initial conditions: *S*(0) = 10927986, *E*(0) = 0.00808 × 1131, *I*(0) = 0.00808 × 482, *A*(0) = 0.00808 × 506, *P*_1_(0) = 0.00808 × 300, *P*_2_(0) = 0.00808 × 400, *Q*(0) = 0.00808 × 647, *R*(0) = 0, *D*(0) = 0. The remaining parameter values are summarised in Table 3 below. Thus, we present numerical simulations of system (1) which was done using the set of parameter values given in Table 3. Some of the parameter values were taken from published literature and some were carefully estimated using the data fitting procedure. The numerical simulations performed in this section are for illustrative purposes only.

### 4.4 Contour plots of parameter on ℛ_0_

Here, we plot the parameters *b, ε* and *θ* on the basic reproduction number, ℛ_0_. In figures 4(a) and 4(b), we observe that an increase in the values of *b* and *ε* results in an decrease in the value of ℛ_0_. Further, the simulation results in figure 4(b) illustrate that prolonging the impact of the effective contact radius in combination with increasing the value of the parameter *b* do not impact the value of ℛ_0_.

**Figure 4.**
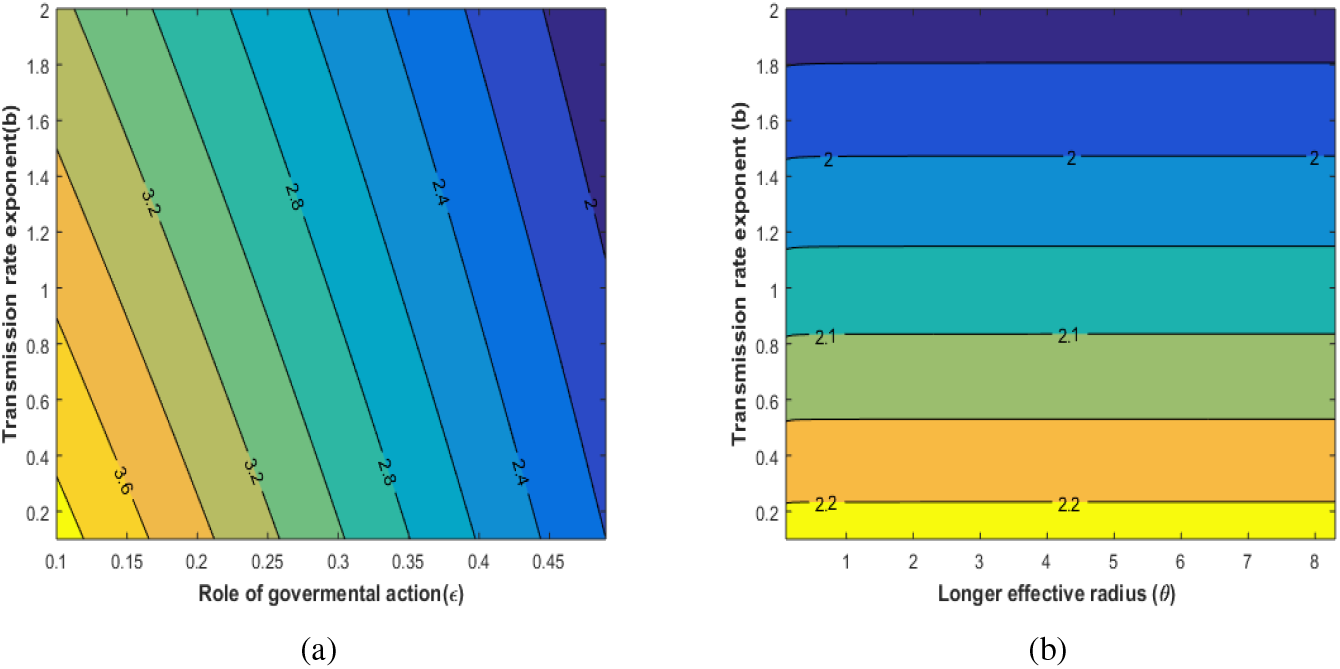
(a) Contour plot of the parameters *b* and *ε* versus ℛ_0_. (b) Contour plot of the parameters *ε* and *θ* versus ℛ_0_. Note that *E* in this figure represents the parameter *ε*, as Matlab do not compile *ε*.

### 4.5 Effects of parameters on ℛ_0_

In this subsection, we show the effects of parameters *r, θ, b* and *ε* on the basic reproduction number. We simulate these parameters as functions of ℛ_0_. In figure 5(a), we observe that an increase in both parameters *r* and *θ* increases the value of ℛ_0_ while figure 5(b) shows that an increase in the values of the parameters *ε* and *b* reduces the value of ℛ_0_. Thus, measures in the form of governmental role actions such as national lockdowns, enforcing effective mask usage, etc are beneficial in curtailing the spread of the disease.

**Figure 5.**
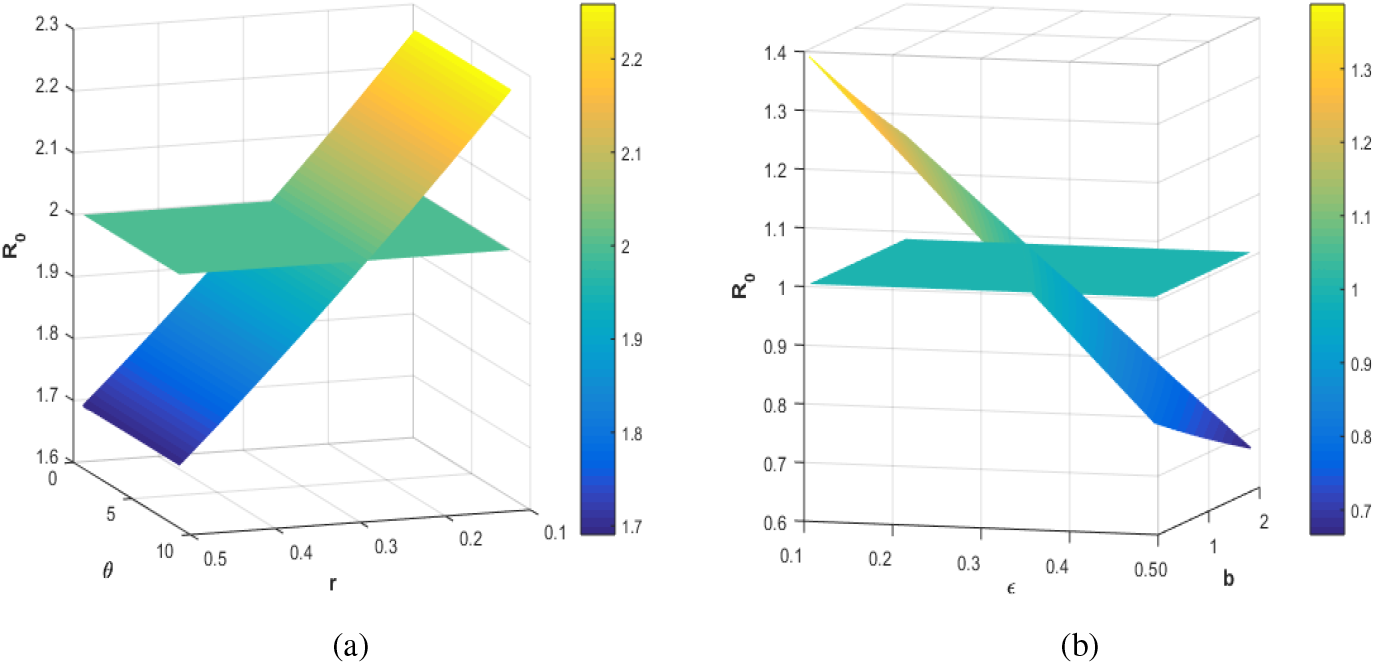
(a) Contour plot of the parameters *r* and *θ* versus ℛ_0_. (b) Contour plot of the parameters *E* and *b* versus ℛ_0_. Note that *E* in this diagram represents the parameter *ε*, as Matlab do not compile *ε*.

### 4.6 Simulation results of parameters on A, I, P_1_ and P_2_

Figures 6-9 illustrate the effect of varying parameters *r, θ, b* and *ε* on the population classes *A*(*t*), *I*(*t*), *P*_1_(*t*) and *P*_2_(*t*). An arrow pointing upwards indicate that as the parameter increases then the number of individuals increases whereas an arrow pointing downwards indicate that as the parameter increases then the number of individuals decreases. It can be observed from figures 6 and 7 that an increase in parameters *r* and *θ* leads to an increase in the number of individuals in compartments *A, I, P*_1_(*t*) and *P*_2_(*t*). Figures 8 and 9 show that an increase in parameters *b* and *ε* results in a decrease in the number of individuals in compartments *A, I, P*_1_(*t*) and *P*_2_(*t*). Thus, efforts targeted at reducing parameters *r* and *θ* and increasing parameters *b* and *ε* will be beneficial in controlling the COVID-19 pandemic. For instance, a reduction in the value of parameter *r* can be achieved through enforcement of measures such as social distancing, curfews e.t.c. which are meant to limit social interactions of individuals in communities.

**Figure 6.**
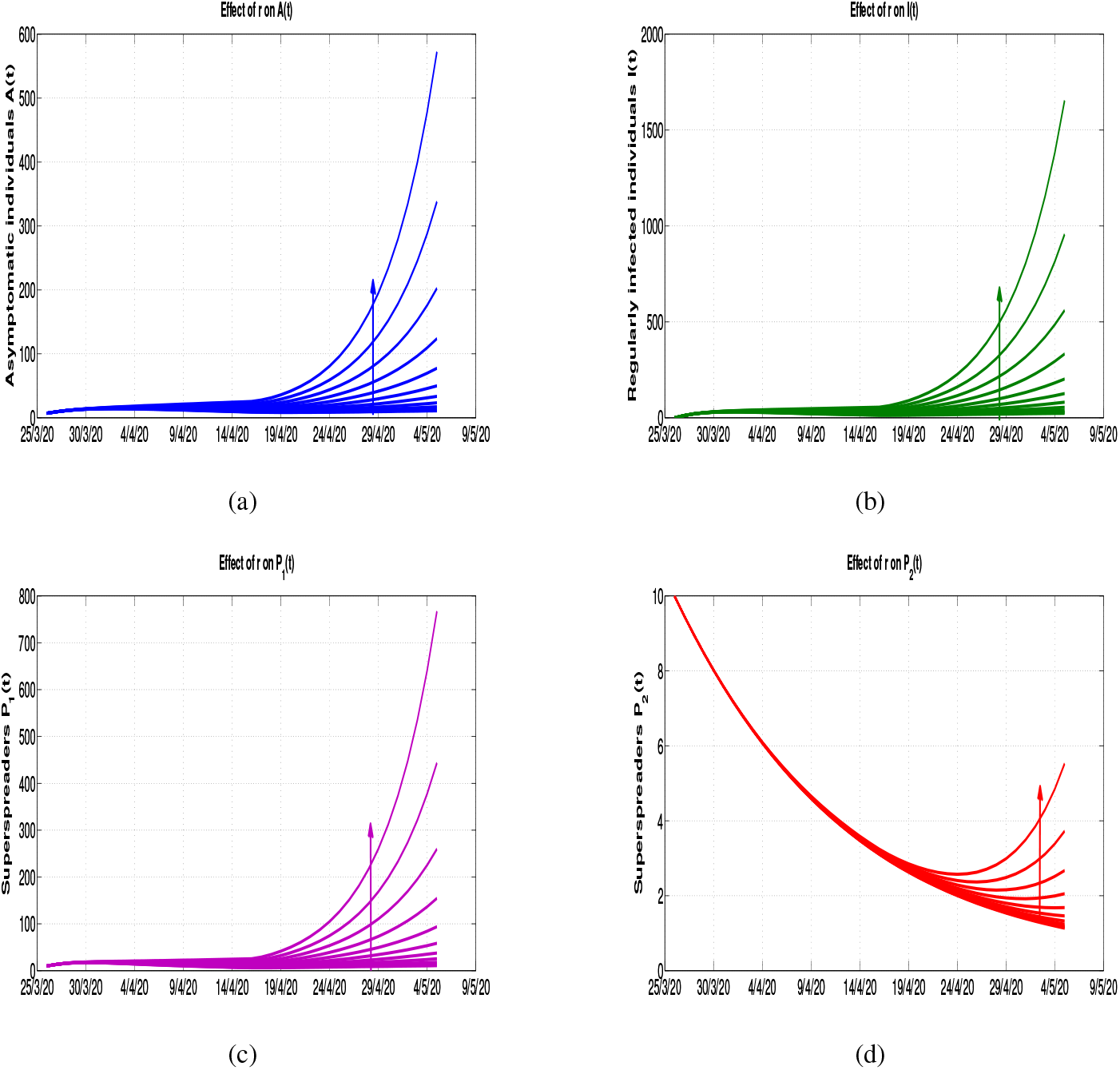
Effects of varying the parameter *r* on the population trajectories of (*a*) asymptomatic individuals *A*(*t*), (*b*) regularly infected individuals *I*(*t*), (*c*) superspreader individuals *P*_1_(*t*) and (*d*) superspreader individuals *P*_2_(*t*), starting from *r* = 0 up to *r* = 2.0 with a step size of 0.2 across all the population compartments.

**Figure 7.**
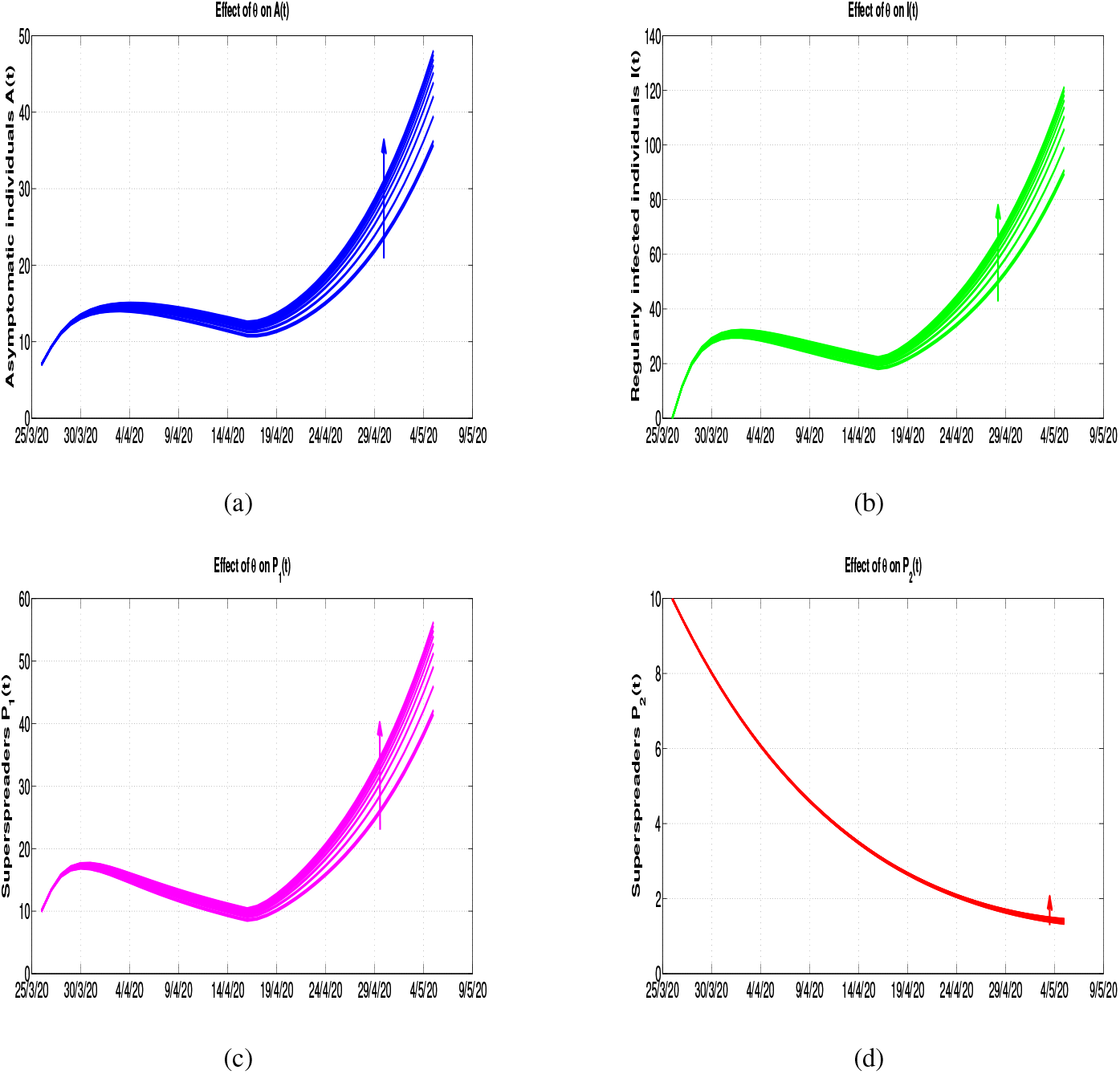
Effects of varying the parameter *θ* on the population trajectories of (*a*) asymptomatic individuals *A*(*t*), (*b*) regularly infected individuals *I*(*t*), (*c*) superspreader individuals *P*_1_(*t*) and (*d*) superspreader individuals *P*_2_(*t*), starting from *θ* = 3 up to *θ* = 9.0 with a step size of 0.3 across all the population compartments.

**Figure 8.**
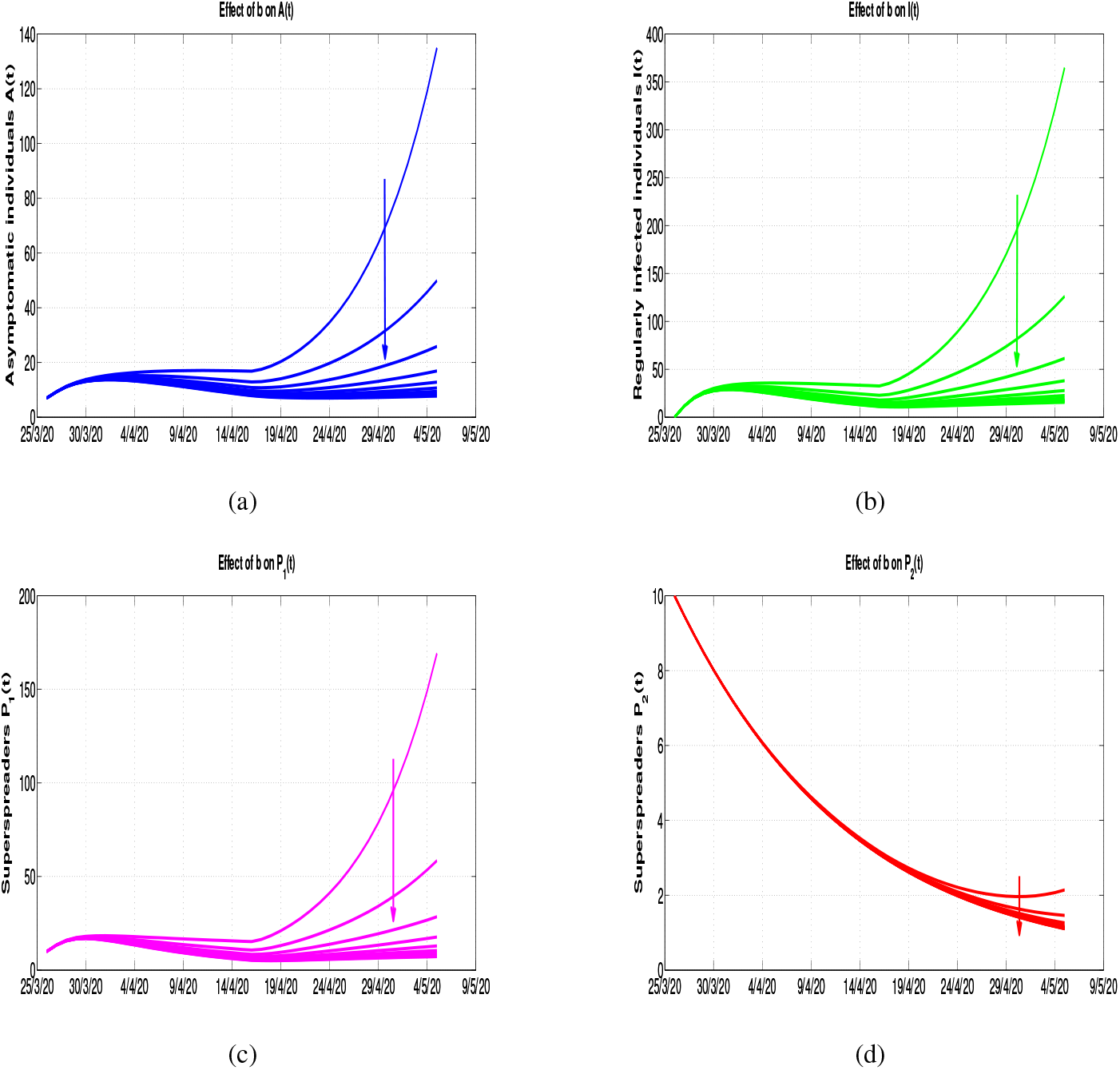
Effects of varying the parameter *b* on the population trajectories of (*a*) asymptomatic individuals *A*(*t*), (*b*) regularly infected individuals *I*(*t*), (*c*) superspreader individuals *P*_1_(*t*) and (*d*) superspreader individuals *P*_2_(*t*), starting from *b* = 1 up to *b* = 10 with a step size of 1.0 across all the population compartments.

**Figure 9.**
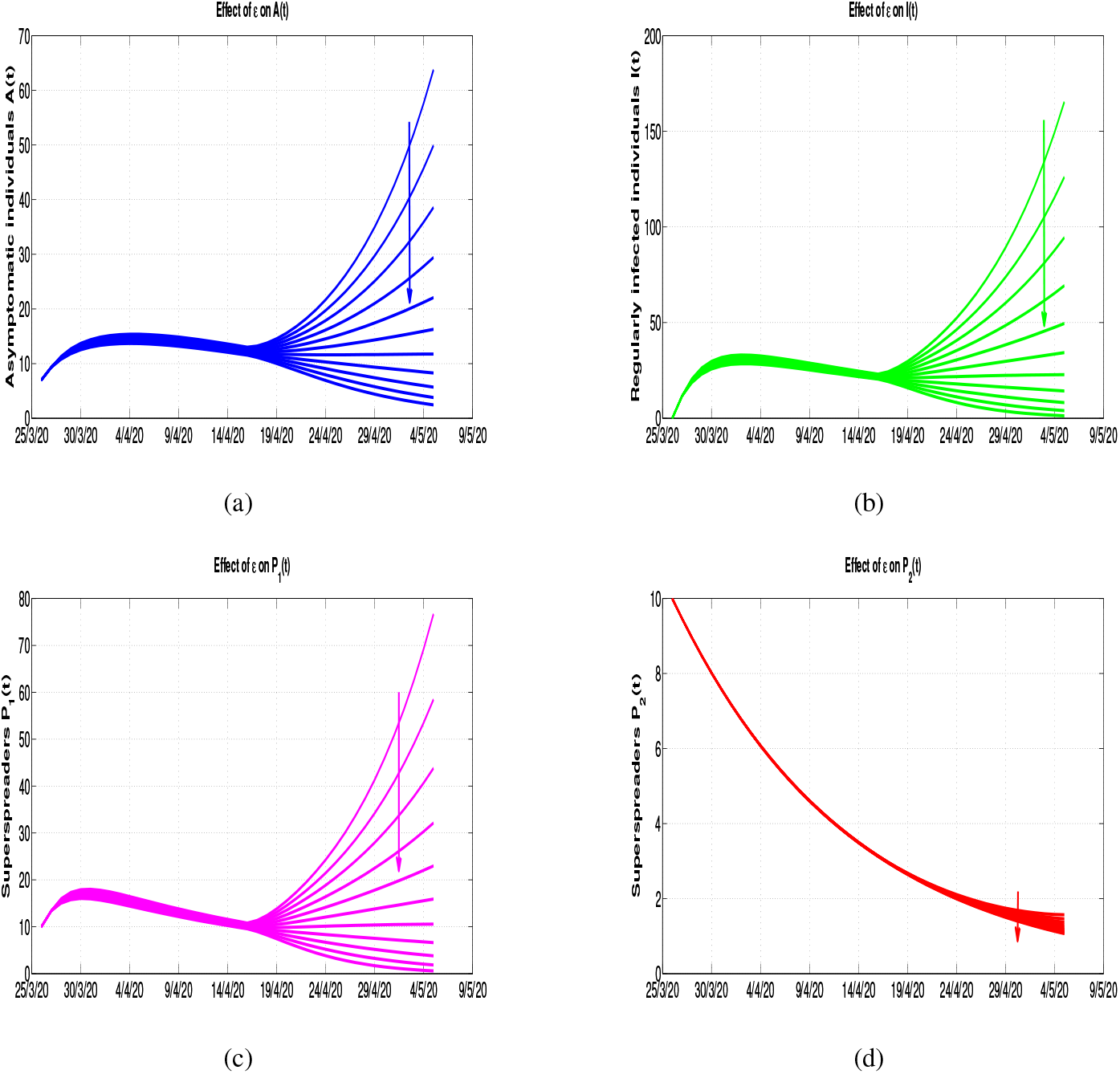
Effects of varying the parameter *ε* on the population trajectories of (*a*) asymptomatic individuals *A*(*t*), (*b*) regularly infected individuals *I*(*t*), (*c*) superspreader individuals *P*_1_(*t*) and (*d*) superspreader individuals *P*_2_(*t*), starting from *ε* = 0 up to *ε* = 1 with a step size of 0.1 across all the population compartments.

## 5 Discussion

A deterministic model incorporating the potential impact of superspreaders on the spread of COVID-19 infection is presented and analysed. The model developed in this paper extends previous works done on SS by taking into account two forms of transmission rate functions for superspreaders based on infectivity level and contact level. Mathematical analysis of the model was done by establishing the positivity and boundedness of the model. The basic reproduction number was calculated and represented in terms of sub reproduction numbers indicating the contributions of each infectious compartment towards new COVID-19 infections.

An application of the SS model was done through fitting the model to epidemiological data for Delhi in India where SS phenomenon was observed for COVID-19. The model was observed to fit well with the data and unknown epidemiological parameter values were estimated within plausible ranges. These parameter values together with some parameter values obtained from the literature were used to carry out numerical simulations. Some few important parameters and important classes of individuals were chosen for numerical results in order to draw important results in line with the objective of the paper. It was noted that an increase of values in parameters *r* and *θ* resulted in an increase in the number of individuals in classes *A*(*t*), *I*(*t*), *P*_1_(*t*) and *P*_2_(*t*) whereas an increase of values in parameters *b* and *ε* resulted in an decrease in the number of individuals in classes *A*(*t*), *I*(*t*), *P*_1_(*t*) and *P*_2_(*t*). Thus, measures meant to increase the values of *b* and *ε* and decrease the values *r* and *θ* will be of great help in the control of COVID-19 in the presence of SS phenomenon. This is also supported by results shown in figure 4. Thus, control measures such as enforcement of social distancing, curfews, wearing of face masks, use of hand sanitizers etc are of great importance in the fight against COVID-19.

The model presented in this paper is not exhaustive. Considering the current intervention strategies taking place in many countries such as vaccination, the model can be extended to study the impact of vaccination. Vital dynamics can as well be incorporated in the model to add realism.

## Data Availability

Delhi COVID-19 data

https://en.wikipedia.org/wiki/COVID-19_pandemic_in_Delhi

## References

Kim Y., Lee S., Chu C. et al. (2016) ‘The Characteristics of Middle Eastern Respiratory Syndrome Coronavirus Transmission Dynamics in South Korea’, Osong Public Health Res Perspect, Vol. 7, No. 1, pp.49–55, http://dx.doi.org/10.1016/j.phrp.2016.01.001.

Hui D.S. (2016) ‘Super-spreading events of MERS-CoV infection’, http://dx.doi.org/10.1016/S0140-6736(16)30828-5

Stein R.A. (2011) ‘Super-spreaders in infectious diseases’, International Journal of Infectious Diseases, Vol. 15, pp.e510–e513, http://dx.doi:10.1016/j.ijid.2010.06.020.

Frieden T.R. and Lee C.T. (2020) ‘Identifying and Interrupting Superspreading Events-Implications for Control of Severe Acute Respiratory Syndrome Coronavirus 2’, Emerging Infectious Diseases, Vol. 26, No. 6, pp.e510-e513, DOI:https://doi.org/eid2606.200495.

Wang D., Hu B., Hu C., Zhu F. et al. (2020) ‘Clinical characteristics of 138 hospitalized patients with 2019 novel coronavirus-infected pneumonia in Wuhan, China’, Epub ahead of print https://doi.org/10.1001/jama.2020.1585.

Wong G., Liu W., Liu Y., Zhou, B. et al. (2015) ‘MERS, SARS, and Ebola: the role of super-spreaders in infectious disease’, Cell Host Microbe, Vol. 18, pp.398–401, https://doi.org/10.1016/j.chom.2015.09.013.

Shen, Z., Ning F., Zhou W., He, X., Lin C., Chin D.P. et al. (2004) ‘Super spreading SARS events, Beijing, 2003’, Emergence Infectious Disease, Vol. 10, pp.256–260, https://doi.org/10.3201/eid1002.030732.

Issarow C.M., Mulder N., Wood R. et al. (2018) ‘Environmental and social factors impacting on epidemic and endemic tuberculosis: a modelling analysis’, Royal Society Open Science, Vol. 5, pp.170726, http://dx.doi.org/10.1098/rsos.170726.

Lau M.S., Dalziel B.D., Funk, S. et al. (2017) ‘Spatial and temporal dynamics of superspreading events in the 2014âŁ”2015 West Africa Ebola epidemic’, Proc Natl Acad Sci USA, Vol. 114, pp.2337–42, https://doi.org/10.1073/pnas.1614595114.

Wallinga J. and Teunis P. (2004) ‘Different epidemic curves for severe acute respiratory syndrome reveal similar impacts of control measures’, American Journal of Epidemiology, Accessed 26 August 2021 https://doi.org/10.1093/aje/kwh255.

Wallinga J. and Teunis P. (2020) ‘Patient 31âŁ™ and South KoreaâŁ™s sudden spike in coronavirus cases’, Aljazeera, Vol. 160, pp.509–16., https://www.aljazeera.com/news/2020/03/31-south-korea-sudden-spike-coronavirus-cases-200303065953841.html.

BBC News (2020) ‘Coronavirus: India âŁ∼super spreaderâŁ™ quarantines 40,000 people.’, Aljazeera, Accessed 28 August 2021, https://www.bbc.co.uk/news/world-asia-india-52061915.

Melsew Y.A., Gambhir M., Cheng A.C., McBryde E.S., Denholm J.T et al. (2020) ‘The role of super-spreading events in Mycobacterium tuberculosis transmission: evidence from contact tracing’, BMC Infectious Diseases, Vol. 19, pp.244 https://doi.org/10.1186/s12879-019-3870-1.

Marineli F., Tsoucalas G., Karamanou M., Androutsos G. (2013) ‘Mary Mallon (1869-1938) and the history of typhoid fever’, Ann Gastroenterol, Vol. 26, pp.132–4 https://doi.org/10.1186/s12879-019-3870-1.

Riley R.L., Mills C.C., O’Grady F., Sultan, L.U. et al. (1962) ‘Infectiousness of air from a tuberculosis ward Ultraviolet irradiation of infected air: comparative infectiousness of different patients’, Am Rev Respiratory Dissease, Vol. 85, pp.511–25 https://doi.org/10.1186/s12879-019-3870-1.

Hethcote H.W. and Yorke J.A. (1984) ‘Modeling Gonorrhea in a Population with a Core Group In: Levin S editor Gonorrhea Transmission Dynamics and Control’, Heidelberg: Springer-Verlag Berlin Heidelberg GmbH, pp.32–48.

Cave E. (2020) ‘COVID-19 Super-spreaders: Definitional Quandaries and Implications’, Asian Bioethics Review, Vol. 12, pp.235–242, https://doi.org/10.1007/s41649-020-00118-2.

Yunhwan K., Hohyung R., Sunmi L. (2018) ‘Agent-Based Modeling for Super-Spreading Events: A Case Study of MERS-CoV Transmission Dynamics in the Republic of Korea’ it International Journal of Environmental Research and Public Health, Vol. 15, pp.2369, doi:10.3390/ijerph15112369.

Kemper J.T. (1980) ‘On the Identification of Super-spreaders for Infectious Disease’ Mathematical Biosciences, Vol. 48, No. 1-2, pp.111–127.

Mkhatshwa T., Mummert A. (2010) ‘Modeling super-spreading events for infectious diseases: Case study SARS’, arXiv 2010, 1007.0908.

Woolhouse M.E., Dye C., Etard J.F., Smith T. et al. (1997) ‘Heterogeneities in the transmission of infectious agents: implications for the design of control programs’, Proc Natl Academy Science USA, Vol. 94, pp.338–342.

Ndairou F., Area I., Nieto J.J., Torres, D.F.M. (2020) ‘Mathematical modeling of COVID-19 transmission dynamics with a case study of Wuhan’, Chaos Solitons and Fractals, Vol. 135,pp.109846. https://doi.org/10.1016/j.chaos.2020.109846.

Maeno Y. (2011) ‘Discovery of a missing disease spreader’, Physica A, Vol. 190, pp.3412–3426, doi:10.1016/j.physa.2011.05.005.

Lloyd-Smith J.O., Schreiber S.J., Kopp P.E., Getz W.M. (2005) ‘Superspreading and the effect of individual variation on disease emergence’, Nature, Vol. 438, pp.355–359.

James A., Pitchford J.W., Plank M.J. (2007) ‘An event-based model of superspreading in epidemics’, Proc. R. Soc. Lond. Ser. B., Vol. 274, pp.741–747.

Garske T. and Rhodes C.J. (2008) ‘The effect of superspreading on epidemic outbreak size distributions’, Journal of Theoretical Biology, Vol. 253, pp.228–237.

Aihaira K., Masuda N., Konno N. (2004) ‘Transmission of severe acute respiratory syndrome in dynamical small-world networks’, Phys Rev E Stat Nonln Soft Matter Phys, Vol. 69, pp.1608–1610.

Zhang D., Wang Y., Zhang Z. (2019) ‘Identifying and quantifying potential super-spreaders in social networks’, Scientific Reports nature reports, Vol. 9, pp.14811. https://doi.org/10.1038/s41598-019-51153-5.

Fujie R., Odagaki T. (2007) ‘Effects of superspreaders in spread of epidemic’, Phys. A Stat. Mech. Its Appl. Vol. 374, pp.843–852.

Yin F., Xia, X., Song N., Zhu L., Wu J. (2020) ‘Quantify the role of superspreaders-opinion leaders-on COVID-19 information propagation in the Chinese Sina-microblog’, PLoS One Vol.15, No. 6, pp.e0234023.

Yang X., Chen L. and Chen J. (1996) ‘Permanence and positive periodic solution for the single-species nonautonomous delay diffusive models’, Computational and Mathematics with Application, Vol.32, No. 4, pp.109–116.

van den Driessche P. and Watmough J. (2002) ‘Reproduction numbers and sub-threshold endemic equilibria for compartmental models of disease transmission’, Mathematical Bioscience, Vol. 180 No.1-2, pp.29–48.

Template:COVID-19 pandemic data/India/Delhi medical cases chart https://en.wikipedia.org/wiki/Template:COVID-19_pandemic_data/India/Delhi_medical_cases_chart Accessed 10/10/2020.

Biswas, S.K., Ghosh, J.K., Sarkar, S. and Ghosh, U., (2020) ‘COVID-19 Pandemic in India: A Mathematical Model Study’, Nonlinear Dynamics, Vol.102, pp.537âŁ”553, https://doi.org/10.1007/s11071-020-05958-z.

https://en.wikipedia.org/wiki/COVID-19_pandemic_in_Delhi, Accessed 27 August 2021.

https://timesofindia.indiatimes.com/india/corona-cases-in-india-30-of-cases-across-india-tied-to-jamaat-event/articleshow/75227980.cms, Accessed 29 August 2021

Balcha A.A., (2020) ‘Curve Fitting and Least Square Analysis to Extrapolate for the Case of COVID-19 Status in Ethiopia’, Advances in Infectious Diseases, Vol. 10 No.3, pp.143.

Lin Q., Zhao S., Gao D. et al. (2020), ‘A conceptual model for the outbreak of Coronavirus disease 2019 (COVID-19) in Wuhan, China with individual reaction and governmental action’, International Journal of Infectious Disease, Vol. 93 No.3, pp.211–6, https://doi.org/10.1016/j.ijid.2020.02.058.

